# Trends in cognitive function before and after myocardial infarction : findings from the China Health and Retirement Longitudinal Study

**DOI:** 10.1101/2023.08.28.23294714

**Authors:** Sijia Zhu, Jing Shang, Jianye Dong, Qingmei Chen, Jianian Hua

## Abstract

**Objectives:** Incident stroke was associated with cognitive dysfunction after stroke and even before stroke. However, cognitive trends prior to myocardial infarction (MI) and the timeline of cognitive decline in a few years following incident MI remain unclear, especially among the Chinese population. We aimed to evaluate whether MI was associated with cognitive change both before and after MI in China.

**Methods:** This cohort study included 11287 participants without baseline heart problems or stroke from the China Health and Retirement Longitudinal Study. The exposure was self-reported MI. The outcomes were scores of cognitive function in five domains, which reflected abilities of episodic memory, visuospatial abilities, orientation, attention and calculation, and global cognition as a summary measure. Linear mixed models explored cognitive function before and after incident MI among the MI participants and the cognitive trends of participants free of MI.

**Results:** During the 7-year follow-up, 421 individuals (3.7% of 11287, mean [SD] age, 60.0 [9.0] years; 59.1% female) experienced MI events. The cognitive scores of participants of both the MI group and the control group without MI declined gradually due to aging. The annual decline rate of the MI group before incident MI was similar to that of the control group during the whole follow-up period. Incident MI was not associated with acute cognitive decline in all five cognitive domains. Moreover, MI did not accelerate the cognitive decline rate after MI compared with the pre-MI cognitive trends. The decline rate of cognitive function after MI was similar to the rate before MI.

**Conclusions:** Different from stroke, participants who had an MI did not show steeper cognitive decline before MI. MI was not associated with acute cognitive decline and accelerated long-term decline after MI. Future studies are needed to learn the mechanisms behind the different patterns of cognitive decline between MI and stroke.

## 1 Introduction

Cognitive impairment and myocardial infarction (MI) are both global health burdens (Yusuf et al., 2001; Wimo et al., 2017). MI is one of the main causes of death in China and Western countries (Thygesen, 2019). Cognitive impairment bears directly on the secondary and tertiary prevention after MI, including healthy lifestyles and medication adherence, and thus increases the all-cause mortality after MI (Artinian et al., 2010; Huber et al., 2019).

Myocardial infarction, together with coronary heart disease (CHD), has previously been linked to cognitive impairment (Gorelick et al., 2011; Kasprzak et al., 2021). Atherosclerosis, the major pathology of CHD, contributes to both clinical and subclinical stroke, which were well-known to cause subsequent cognitive impairment (de la Torre, 2002; Koton et al., 2022). Heart failure and low perfusion after MI were also reported to have negative effects on cognitive function (Wolters et al., 2018; Vishwanath et al., 2022). However, several knowledge gaps are largely unknown. Firstly, whether the cognitive impairment is due to the cardiovascular event itself or to the cardiovascular risk factors (e.g., hypertension, diabetes) which were known to contribute to both cardiovascular event and cognitive impairment remains unelucidated (Gottesman and Johansen, 2021). The cognitive decline caused by hypertension or diabetes is slow and progressive. If cardiovascular risk factors the main indicator of cognitive decline, patients’ cognitive function may have already declined even before MI onset (Zheng et al., 2019). Nevertheless, most studies assessed cognitive function after the onset of heart disease. For example, clinical doctors could only achieve the cognitive data during hospitalization or after discharge. Surgeries are interested in the risk of dementia after bypass graft surgery, but hard to ascertain the cognitive status before the surgery (Greaves et al., 2020). Cognitive function before MI onset, an important confounder, is impossible to obtain. Second, the previous studies learning the risk of cognitive impairment of CHD patients yield mixed results (Schievink et al., 2017). A meta-analysis identified 28 articles and showed a pooling result that CHD increased the risk of all-cause cognitive impairment (RR = 1.27), while 13 of them reported insignificant association (Liang et al., 2021). Cognitive impairment is initiated from and characterized by decline in cognitive function. In this study, we explored whether cognitive decline had existed before MI and the trajectories of cognitive decline after MI, and therefore provided a better understanding of the effect of MI on cognitive health. Third, previous studies learning the cognitive trajectories before and after CHD are from the UK and America, evidence from the Chinese population is scarce (Xie et al., 2019; Johansen et al., 2023).

The current study aimed to explore cognitive trends both before and after incident MI using a large nationally representative sample from the China Health and Retirement Longitudinal Study (CHARLS).

## 2 Materials and methods

### 2.1 Study Population

Data was obtained from waves 1 to 4 of the CHARLS, an ongoing prospective research which recruited a total of 17,708 participants across 28 provinces in China between 2011 and 2012 at baseline (Supplementary Text 1). The follow-up surveys were conducted at 2-to 3-year intervals till 2018. The comprehensive study design has already been covered elsewhere (Zhao et al., 2012).

Figure 1 presents the sample selection process. 4,663 participants were excluded at baseline because of being under 45 years old (n = 352), missing baseline covariates (n = 71) or baseline cognitive tests (n = 1,668), having a history of memory-related disease (n = 562), brain damage (n = 392), stroke (n = 562), or heart disease (n = 1,970). Furthermore, 1,758 of the remaining individuals were disqualified due to loss to follow-up (not completing at least one cognitive assessment from waves 2 to 4, n = 1,166) or experienced a stroke during follow-up (n = 610). Finally, 11,287 individuals were enrolled in the current study, including 421 participants who had an MI during the follow-up and 10,866 participants without MI.

**Figure 1.**
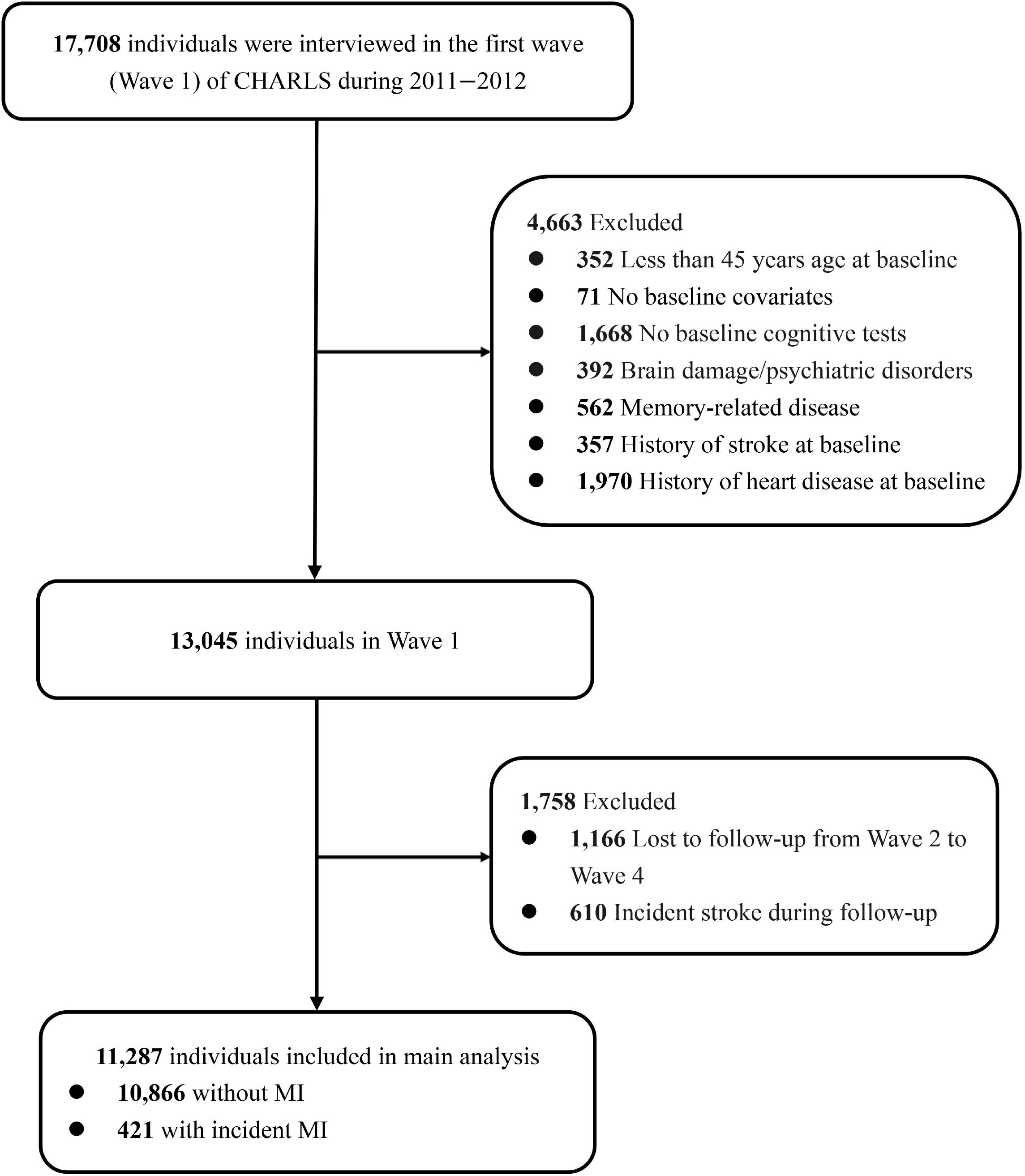
Flow chart of study participants.

### 2.2 Assessment of cognitive function

CHARLS collected participants’ cognitive function at each visit (Wave 1 to Wave 4). Four cognitive domains were assessed by different tests and the detailed methods are as follows: (1) Episodic memory: Word recall test was utilized to estimate both immediate and delayed memory. The investigator read 10 words while participants should recall these words immediately and after 5 minutes. The total scores were 10. (2) Visuospatial ability: Individuals were shown two overlapping pentagons and people who successfully replicated the drawing could get one point in the figure drawing test. (3) Orientation ability: It is assessed by answering the date (day, month, year), week, and season. The score ranged from 0-5. (4) Calculation and attention ability: Participants were asked to perform five consecutive subtractions of 7 from 100 and each correct answer got 1 point (total score: 5). The effectiveness and dependability of these cognitive tests have been clearly demonstrated in the past investigations (Zhao et al., 2014; Meng et al., 2019; Hua et al., 2023).

In order to acquire a more intuitive comparison across different cognitive tasks, z scores were generated by the mean and SD of baseline scores. The z-score of global cognition, a summary of performance on the above four cognitive domains, was calculated by adding the z-scores of the four cognitive domains and then normalizing them to baseline in the same manner. Hence, a z-score of -1 indicates a cognitive score being 1 SD below the average baseline score.

### 2.3 Assessment of incident MI

In Wave 1, participants were asked whether they had been told by a doctor that they had heart problems, including MI, angina, CHD, congestive heart failure, and other heart problems (Zhao et al., 2014). No information about the type of heart problem is available. Those with heart problems at Wave 1 were excluded according to our selection criteria. From Wave 2 to Wave 4, MI was based on self or proxy reports of physician diagnosis. However, other types of heart disease were not recorded. Some participants reported the year of MI. The time of MI of other participants was defined as the midpoint of the last wave without MI and the first wave reporting MI.

### 2.4 Covariates

Details of the covariates are described in Supplementary Text 2 (Cheng and Chan, 2005; Zhao et al., 2014). Covariates were chosen based on the association with cognitive function and MI, which included age, sex, educational level, marital status, area of residence, smoking, drinking, number of instrumental activities of daily living (IADLs), depression, hypertension, diabetes, high total cholesterol, lung diseases, and cancer (Zheng et al., 2019).

### 2.5 Statistical analysis

Baseline characteristics between participants with and without incident MI, and between participants included and excluded due to loss of follow-up were compared. Our primary analysis was a two-level linear model (Supplementary Figure 1). The model accommodated the repeated measurements of cognitive function over time and calculated the effect of MI and other covariates on the trajectories of cognitive function (Laird and Ware, 1982; West et al., 2007; Bethany A. Bell, 2013; Berglund, 2018). Fixed effects were fitted for intercept, baseline covariates, group (the group with incident MI and the control group without MI), time (years since baseline), ‘group*time interaction’, a time-varying MI status (whether or not had a history of MI at the current wave), and ‘group*MI-status*years-after-MI’ interaction. Random effects were fitted for intercept and time (years since baseline and years after MI). In subgroup analysis, we restricted the analysis to those with incident MI during the follow-up. Interaction terms between covariates and cognitive trajectories were added to explore potential risk factors for cognitive change after MI. CHARLS did not measure body mass index (BMI) in Wave 1. In sensitivity analysis, we further adjusted for BMI which was recorded in Wave 2 or Wave 3. Participants who had a stroke during follow-up were included to evaluate whether the cognitive change was caused by stroke. Since the included participants tended to be healthier than the excluded participants, our primary analysis was further restricted to individuals who received cognitive assessments in all waves to achieve more accurate results among the healthier participants.

Statistical analyses were performed from April 2023 to July 2023 using SAS version 9.4 (SAS Institute Inc.). A two-sided *P* < 0.05 was considered statistically significant.

## 3 Results

### 3.1 Baseline characteristics and cognitive tests

During the 7-year follow-up, 421 individuals (3.7% of 11287 included individuals, mean [SD] age, 60.0 [9.0] years; 59.1% female) experienced incident MI. Baseline characteristics according to incident MI are shown in Table 1. The number of incident MI between each wave is shown in Supplementary Table 1. From waves 1 to 4, the available cognitive assessments were 421, 386, 383, and 300 in the MI group, and 10,866, 9,694, 9,420, and 7,511 in the control group (Supplementary Table 2). Characteristics of 1,148 (8.9%) participants who lost to follow-up are shown in Supplementary Table 3.

**Table 1.** Baseline participant characteristics according to MI.

|  | Without MI<br>(n=10866) | Incident MI<br>(n=421) | *p* value<sup>*</sup> | Adjusted *p* value<sup>†</sup> |
| --- | --- | --- | --- | --- |
| Age, y | 58.2 (9.2) | 60.0 (9.0) | <0.001 | None |
| IADLs, No. | 0.1 (0.6) | 0.2 (0.7) | 0.017 | 0.091 |
| Males | 5252 (48.3) | 172 (40.9) | 0.003 | None |
| Education |  |  | 0.499 | 0.014 |
| Illiterate | 2891 (26.6) | 118 (28.0) |  |  |
| Primary school | 4345 (40.0) | 156 (37.1) |  |  |
| Middle school | 2307 (21.2) | 99 (23.5) |  |  |
| High school and above | 1323 (12.1) | 48 (11.4) |  |  |
| Married | 9713 (88.8) | 368 (87.4) | 0.510 | 0.645 |
| Living area |  |  | 0.019 | 0.017 |
| Urban | 2128 (19.6) | 102 (24.2) |  |  |
| Rural | 8738 (80.4) | 319 (75.6) |  |  |
| Current smoking | 4254 (39.2) | 147 (34.9) | 0.081 | 0.720 |
| Current drinking | 3161 (26.8) | 88 (20.0) | 0.002 | 0.059 |
| Depression | 2075 (19.1) | 111 (26.7) | <0.001 | 0.003 |
| Hypertension | 2778 (25.6) | 173 (41.1) | <0.001 | <0.001 |
| Diabetes | 709 (5.8) | 51 (11.8) | <0.001 | <0.001 |
| High total cholesterol | 1460 (13.4) | 89 (21.1) | <0.001 | <0.001 |
| Lung Diseases | 903 (8.3) | 61 (14.5) | <0.001 | <0.001 |
| Cancer | 88 (0.8) | 5 (1.2) | 0.394 | 0.487 |

<sup>\*</sup>Calculated using *t*-test for continuous covariates and $\chi^2$ or Fisher test for categorical covariates.
<sup>†</sup>Calculated using generalized linear models for continuous covariates and logistic regression for categorical covariates after adjustment for baseline age and sex.

### 3.2 Difference between the MI group and the control group during the pre-MI period

For the control group without MI, scores of all cognitive domains declined during the whole follow-up period (Table 2). This is mainly due to aging. The baseline cognitive scores of the MI group were not different from the baseline scores of the control group. Meanwhile, the change rate (slope) of cognitive scores among the MI group during the pre-MI period were similar to that among the control group from baseline to the end of follow-up. In conclusion, the cognitive function of the MI group declined gradually before MI onset but was not weaker than the cognitive function of those without MI.

**Table 2.** Difference in cognitive trends between the MI group and the control group before MI^a,b^.

|  | Cognitive slope of control group |  | Difference in baseline cognitive scores |  | Difference in cognitive slope before MI |  |
| --- | --- | --- | --- | --- | --- | --- |
| | $\beta$ (95% CI) | *p* | $\beta$ (95% CI) | *p* | $\beta$ (95% CI) | *p* |
| Global cognition | -0.036 (-0.038, -0.033) | <0.001 | 0.074 (-0.007, 0.154) | 0.073 | -0.001 (-0.028, 0.025) | 0.920 |
| Episodic memory | -0.007 (-0.010, -0.003) | <0.001 | 0.027 (-0.069, 0.123) | 0.582 | 0.000 (-0.032, 0.032) | 0.990 |
| Visuospatial abilities | -0.037 (-0.041, -0.034) | <0.001 | 0.016 (-0.079, 0.111) | 0.742 | 0.011 (-0.022, 0.043) | 0.520 |
| Orientation | -0.036 (-0.038, -0.033) | <0.001 | 0.032 (-0.048, 0.112) | 0.428 | -0.008 (-0.035, 0.020) | 0.581 |
| Attention and calculation | -0.052 (-0.055, -0.049) | <0.001 | 0.070 (-0.021, 0.162) | 0.129 | 0.015 (-0.014, 0.045) | 0.306 |

<sup>a</sup>Adjusted for baseline age, sex, educational level, marital status, area of residence, smoking, drinking, number of instrumental activities of daily living (IADLs), depression, hypertension, diabetes, high total cholesterol, lung diseases, and cancer.
<sup>b</sup>All cognitive values are z-score transformed.

### 3.3 Difference between pre-MI and post-MI cognitive trends

As shown in Table 3, participants did not experience an acute cognitive change at the time of MI. The cognitive slope in the long time after MI was not statistically different from the pre-MI cognitive slope. Therefore, the cognitive decline rate post-MI was comparable with the pre-MI rate. In most of the subgroups, incident MI was not associated with acute cognitive change at the time of MI or changes in cognitive slope in the years following MI (Figure 2 and Supplementary Figure 2). The cognitive function of MI participants with lung disease acutely increased at the time of MI in domains of global cognition and visuospatial abilities. In all other subgroups, there was no statistically significant cognitive change after MI.

**Table 3.** Changes in cognitive trends after MI compared to the pre-MI period^a,b^.

|  | Acute cognitive change after MI |  | Change in cognitive slope after MI |  |
| --- | --- | --- | --- | --- |
| | $\beta$ (95% CI) | *p* | $\beta$ (95% CI) | *p* |
| Global cognition | 0.013 (-0.085, 0.111) | 0.792 | -0.007 (-0.040, 0.027) | 0.691 |
| Episodic memory | 0.023 (-0.096, 0.142) | 0.703 | 0.003 (-0.038, 0.043) | 0.898 |
| Visuospatial abilities | -0.032 (-0.164, 0.100) | 0.638 | -0.021 (-0.065, 0.024) | 0.355 |
| Orientation | 0.011 (-0.099, 0.120) | 0.851 | 0.008 (-0.026, 0.043) | 0.635 |
| Attention and calculation | 0.020 (-0.099, 0.139) | 0.741 | -0.026 (-0.063, 0.011) | 0.172 |

<sup>a</sup>Adjusted for baseline age, sex, educational level, marital status, area of residence, smoking, drinking, number of instrumental activities of daily living (IADLs), depression, hypertension, diabetes, high total cholesterol, lung diseases, and cancer.
<sup>b</sup>All cognitive values are z-score transformed.

**Figure 2.**
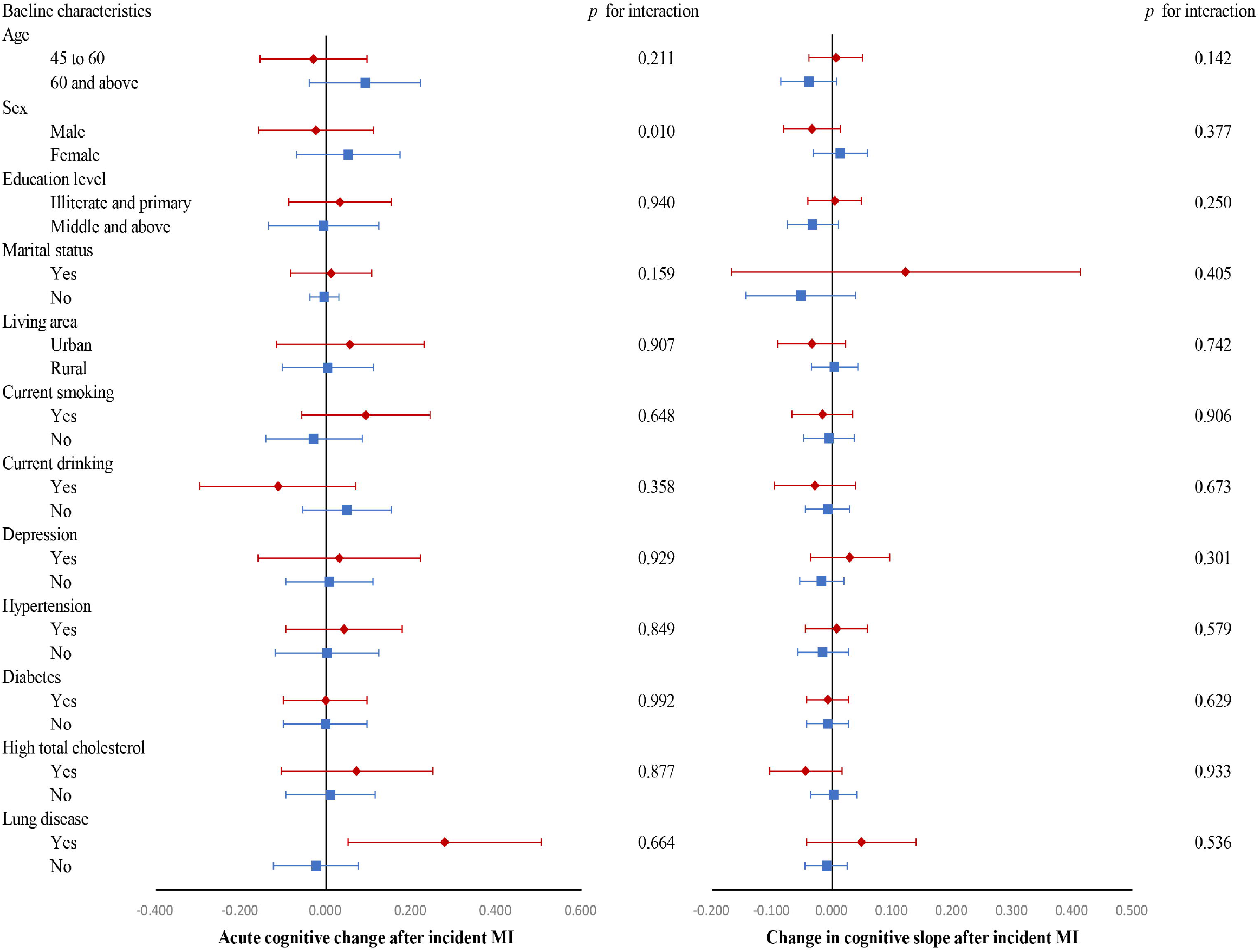
Changes in global cognitive trajectories after MI compared to the pre-MI period according to subgroups. Adjusted for baseline age, sex, educational level, marital status, area of residence, smoking, drinking, number of instrumental activities of daily living (IADLs), depression, hypertension, diabetes, high total cholesterol, lung diseases, and cancer. All cognitive values are z-score transformed.

### 3.4 Sensitivity analysis

To assess the stability of our results, after restricting the study to participants who had cognitive data in all four waves, the MI group had higher baseline scores in attention and calculation domain but still had non-significant acute cognitive change or post-MI cognitive slope change, which is in consistence with our results. In other sensitivity analysis, the MI group showed similar pre-MI cognitive slope compared with the control group. MI group also did not show acute cognitive change or changes in cognitive slope after MI (Supplementary Table 4-6).

## 4 Discussion

Among 11287 participants in this community-based cohort study, the MI group did not show impaired cognitive function before the onset of MI. MI was not associated with acute cognitive decrement at the time of MI or accelerated cognitive deterioration in the long time following MI.

At least five studies had learned the timeline of cognitive decline before and after stroke (Levine et al., 2015; Zheng et al., 2019; Eng et al., 2021; Heshmatollah et al., 2021; Hua et al., 2023). Our previous CHARLS study discovered that stroke was associated with acute cognitive decline in the short term after stroke and accelerated cognitive decline in the future among the Chinese population (Hua et al., 2023). Studies learning the European population further indicated that stroke participants had shown faster cognitive decline even before stroke, suggesting cognitive decline as a predictor for stroke onset (Zheng et al., 2019; Heshmatollah et al., 2021). Previous studies connected the pre-disease cognitive decline to the effect of vascular risk factors which is hard to be adjusted in the statistical models (Whalley et al., 2006; Zheng et al., 2019). Considering that MI and stroke are both vascular diseases, it is tempting to postulate that MI patients had cognitive disadvantages before MI and experienced steeper annual cognitive decline after MI. To the best of our knowledge, two articles have learned the trends in cognition before and after CHD. *Xie* et al. 2019 analyzed participants in the UK from the English Longitudinal Study of Ageing (ELSA) and unveiled that CHD was not associated with pre-CHD cognitive disadvantage or acute cognitive change (Xie et al., 2019). By pooling six cohort studies from the US, *Johansen* et al. 2023 also revealed that MI patients had similar cognitive trajectories before MI compared with the without-MI participants and did not suffer from acute cognitive decline (Johansen et al., 2023). These results are in accordance with our findings. However, our results in cognitive slopes in years after MI were different from the above two articles. Xie et al. 2019 disclosed that the decline rate accelerated after incident CHD in domains of global cognition, memory, and orientation, but not semantic fluency. *Johansen* et al. 2023 narrated that the first MI was correlated with faster declines in memory, executive function, and global cognition over time and that the results differed by race and sex. Notably, *Whilock* et al. 2021 reported that the cognitive slope before and after coronary artery bypass grafting (CABG) and percutaneous coronary intervention (PCI) were similar in the Health and Retirement Study (HRS) in US (Whitlock et al., 2021).

Why the decline rate of cognitive function of the stroke population was steeper than that of the without-stroke population during the pre-stroke period, but the decline rate of the MI group was similar to the rate of the without-MI group during the pre-MI period? Although stroke and MI mainly result from atherosclerosis and atrial fibrillation, the pre-disease cognitive patterns are different. Let us make a bold assumption. The pre-stroke cognitive disadvantage is not because of vascular risk factors (e.g., diabetes, smoking, or atherosclerosis) but because of the intracranial damage (e.g., subclinical brain infarcts and white matter hyperintensity) (Dhamoon et al., 2018). If our hypothesis were correct, performing head magnetic resonance imaging (MRI) might predict pre-stroke cognitive decline and onset of stroke.

Our study added to prior stroke research which learned the timeline of cognitive decline after stroke. It is widely reported that the incident rate of dementia within 6 months after a stroke was much higher than the incident rate several years after stroke (Pendlebury et al., 2019). The concept, named ‘acute cognitive change/decline’ in the linear mixed model, could explain this phenomenon, and it is obviously caused by the strike of acute illness inside the brain (Baron et al., 1981; Garcia-Alloza et al., 2011). The absence of acute cognitive change after MI supported that the effect of MI on the brain happened gradually rather than suddenly.

This is one of the largest important cohort studies reporting the cognitive trends before and after MI. Another advantage of our study is the study population. Chinese participants have different economic and educational levels, which would lead to different patterns of cognitive decline after the illness (Levine et al., 2018). However, several limitations need to be acknowledged. Firstly, CHARLS did not differentiate the type of heart problems in the first wave and only recorded MI in the latter waves. This prevented us from considering subgroups of atrial fibrillation, angina, or asymptomatic CHD. We were also unable to investigate the prognosis of different MI treatments (conservative internal medical treatment, PCI, and CABG). Fortunately, recording all kinds of heart problems enabled a strict exclusion criterion during sample selection. Second, the sensitivity and specificity of self-reported MI in CHARLS are unknown. Part of the participants would mistake unstable angina or arrhythmias for MI (Rosamond et al., 1995). Lower positive predictive values for self-reported MI would underestimate the negative effect of MI on cognition. Third, our follow-up period is relatively short. The results could only reflect the cognitive trends during several years after MI. As the follow-up survey of CHALRS is still continuing, future studies could re-examine the cognitive trends after MI.

Cognitive function of the MI group was not poorer than the without-MI group before MI onset. MI was not associated with acute cognitive decline in the short term after MI or accelerated cognitive decline in the years following MI. The cognitive pattern before an incident MI conflicted with that before an incident stroke. Shared cardiovascular risk factors for MI and stroke could not explain the pre-stroke cognitive discrepancy. The mechanism underlying the difference is unknown and warrants future study.

## 6 Author Contributions

JH and SZ conceived the study and designed the statistical analyses. JH organized the database. JH, SZ and JD did the statistical analyses and prepared the draft of the manuscript. JH, QC, and JS substantively revised the manuscript. All authors contributed to the interpretation of data. The corresponding authors attest that all listed authors meet authorship criteria and that no others meeting the criteria have been omitted. All authors read and approved the final manuscript.

## 8 Acknowledgments

We appreciated the China Center for Economic Research and the National School of Development of Peking University for providing the data.

## 9 Data Availability Statement

Data used in this manuscript from the China Health and Retirement Longitudinal Study (CHARLS). We applied permission for the data access (http://charls.pku.edu.cn/zh-CN) and obtained access to use it. Prof. Yaohui Zhao (National School of Development of Peking University), John Strauss (University of Southern California), and Gonghuan Yang (Chinese Center for Disease Control and Prevention) are the principal investigators. Requests to access these data can also be directed to Jianian Hua.

## 10 List of abbreviations

BMI: body mass index;
CABG: coronary artery bypass grafting;
CHARLS: China Health and Retirement Longitudinal Study;
CHD: coronary heart disease;
ELSA: English Longitudinal Study of Ageing;
HbA_1c_: hemoglobin A_1c_;
HRS: Health and Retirement Study;
IADLs: instrumental activities of daily living;
MI: myocardial infarction;
MRI: magnetic resonance imaging;
PCI: percutaneous coronary intervention.

