## Supplementary Material for "Trends in cognitive function before and after myocardial infarction : findings from the China Health and Retirement Longitudinal Study"

### 1 Supplementary Text

#### 1.1 Text S1. Ethics approval

Ethical approval for all the CHARLS waves was granted from the Institutional Review Board at Peking University. The IRB approval number for the main household survey, including anthropometrics, is IRB00001052-11015; the IRB approval number for biomarker collection is IRB00001052-11014. During the fieldwork, each respondent who agreed to participate in the survey was asked to sign two copies of informed consent; one copy was kept in the CHARLS office, which was also scanned and saved in PDF format. Four separate consents were obtained: one for the main fieldwork, one for the non-blood biomarkers and one for the taking of the blood samples, and another for storage of blood for future analyses.

#### 1.2 Text S2. Methods

#### Covariates

Educational attainment was divided into illiterate, primary school, middle school, and high school and above. Marital status was dichotomized into married and other status. Body mass index (BMI) was categorized into underweight ($<$18.5 kg/m2), normal weight (18.5–23.9 kg/m2), and overweight or obese (≥24 kg/m2). Hypertension was defined as self-reported diagnosis, use of antihypertensive medication, or the mean of three systolic/diastolic blood pressure measurements ≥140/90 mm Hg. High total cholesterol was defined as self-reported diagnosis, use of lipid-lowering medication, or serum total cholesterol ≥ 240mg/dl. Depressive symptoms were measured using the 10-item Centre for Epidemiologic Studies Short Depression Scale, with depression defined as a score of ≥12 from a total score of 0 to 30.

### 2 Supplementary Figures

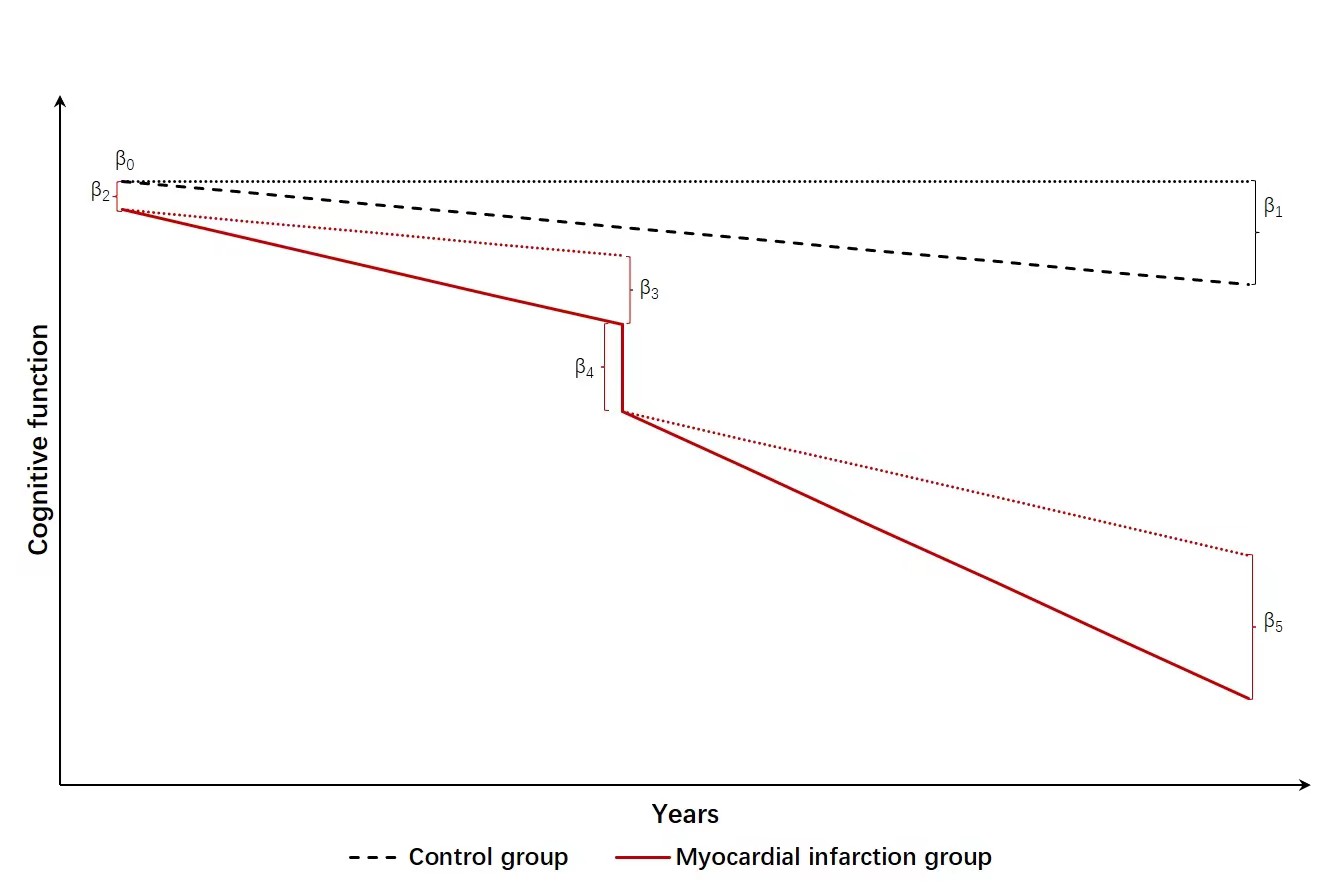

**Supplementary Figure 1. The conceptual model of our study.** Time on the x-axis is the years from the date of the first cognitive test. Y-axis is the cognitive function. The black dashed line represents the possible trajectory of the control group without myocardial infarction (MI). We hypothesized their cognitive function declined annually due to aging. The cognitive trajectories of the MI group (red lines) consisted of the trajectories before MI, an acute cognitive decline/change at the time of MI, and an accelerated decline (change in slope) after MI. $\beta$_0_: The predicted cognition of the control group at baseline (time t=0). $\beta$_1_: The average slope of cognition of the entire control group. $\beta$_2_: The difference in cognition at baseline in the MI group compared to the control group. $\beta$_3_: The difference in slope of the MI group compared to the control group before MI. $\beta$_4_: The ‘acute cognitive change/decline’ at the MI point among the MI group, measured as the first predicted post-MI cognitive score minus the last predicted pre-MI cognitive score. $\beta$_5_: The change in slope in the post-MI period compared to the pre-MI period. After MI, we hypothesized the cognitive decline rate was combined with the pre-MI decline rate and an accelerated decline caused by MI. We assumed MI affects cognition in all years after stroke.

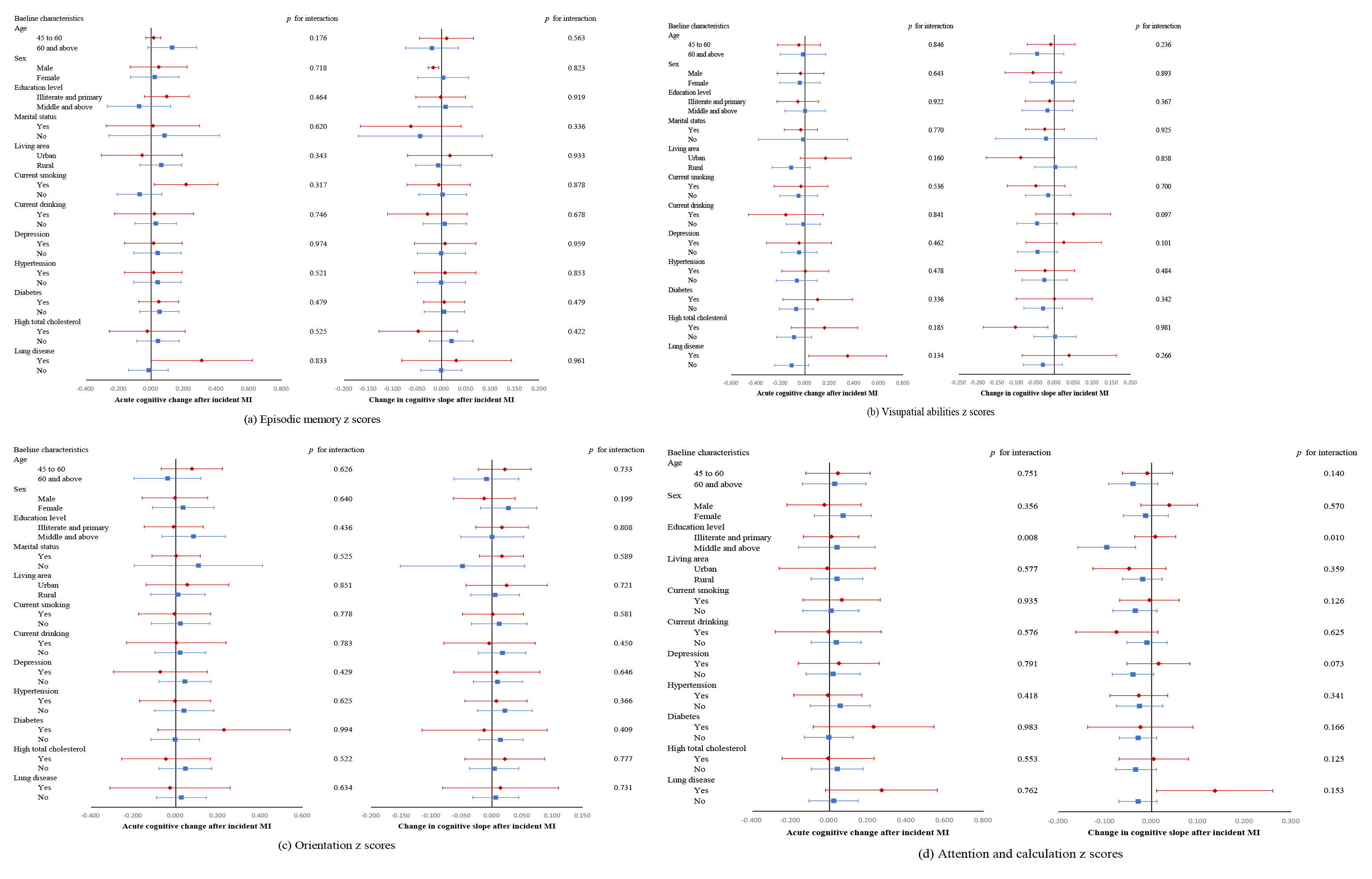

Supplementary Figure 2. **Changes in cognitive trajectories after MI compared to the pre-MI period according to subgroups.** Adjusted for baseline age, sex, educational level, marital status, area of residence, smoking, drinking, number of instrumental activities of daily living (IADLs), depression, hypertension, diabetes, high total cholesterol, lung diseases, and cancer. All cognitive values are z-score transformed.

### 3 Supplementary Tables

#### 3.1 Supplementary Table 1. Number of incident MI between each wave

| Diagnosis time | Number of  incident MI |
| --- | --- |
| 0 to 2 years | 149 |
| 2 to 4 years | 177 |
| 4 to 7 years | 93 |

#### 3.2 Supplementary Table 2. Number of available cognition measurements in each wave

|  | Wave 1 | Wave 2 | Wave 3 | Wave 4 |
| --- | --- | --- | --- | --- |
| Without-MI | 10866 (100%) | 9694 (89.2%) | 9420 (86.7%) | 7511 (69.1%) |
| Incident MI | 421 (100%) | 386 (91.7%) | 383 (90.9%) | 300 (71.3%) |

#### 3.3 Supplementary Table 3. Comparison of baseline characteristics between participants included (n=11287) and excluded due to loss to follow-up (n=1148)^a^

|  | Included | Excluded | *p* value^a^ |
| --- | --- | --- | --- |
|  | (n = 11287) | (n = 1148) |  |
| Continuous variables, mean (SD) |  | | |
| Age | 58.3 (9.3) | 63.4 (12.7) | <0.001 |
| Number of IADLs | 0.1 (0.6) | 0.5 (1.2) | <0.001 |
| Episodic memory | 3.3 (1.9) | 2.7 (2.2) | <0.001 |
| Visuospatial ability | 0.7 0.5) | 0.6 0.5) | <0.001 |
| Orientation | 3.8 1.4) | 3.5 (1.7) | <0.001 |
| Calculation and attention | 2.8 (2.0) | 2.5 (2.0 | <0.001 |
| Categorical variables, n (%) |  | | |
| Males | 5424 (48.0) | 573 (49.9) | 0.230 |
| Education |  |  | <0.001 |
| Illiterate | 3009 (26.6) | 386 (33.6) |  |
| Primary school | 4501 (39.8) | 374 (32.6) |  |
| Middle school | 2406 (21.3) | 196 (17.1) |  |
| High school and above | 1371 (12.2) | 192 (16.7) |  |
| Married | 10081 (89.3) | 897 (78.1) | <0.001 |
| Livubg area |  |  | <0.001 |
| Urban | 2230 (19.8) | 486 (42.3) |  |
| Rural | 9057 (80.2) | 662 (57.7) |  |
| Current smoking | 4401 (39.0) | 477 (41.6) | 0.091 |
| Current drinking | 2991 (26.5) | 258 (22.5) | 0.003 |
| Depression | 2189 (19.4) | 237 (20.7) | 0.304 |
| Hypertension | 2951 (26.1) | 380 (33.1) | <0.001 |
| High total cholesterol | 1549 (13.7) | 139 (12.1) | 0.128 |
| Diabetes | 680 (6.0) | 77 (6.7) | 0.357 |
| Lung Diseases | 964 (8.5) | 118 (10.3) | 0.047 |
| Cancer | 93 (0.8) | 22 (1.9) | <0.001 |

^a^Calculated using ANOVA for continuous covariates and χ^2^ test for categorical covariates.

#### 3.4 Supplementary Table 4. Trajectories of cognitive z scores among all participants over time after including participants who had a stroke during follow-up (464 in the MI group and 11416 in the control group)^a,b^

|  | Global cognition |  | Episodic Memory |  | Visuospatial ability |  | Orientation |  | Attention and calculation |  |
| --- | --- | --- | --- | --- | --- | --- | --- | --- | --- | --- |
|  | β (95% CI) | *p* | β (95% CI) | *p* | β (95% CI) | *p* | β (95% CI) | *p* | β (95% CI) | *p* |
| Variables |  |  |  |  |  |  |  |  |  |  |
| Baseline age | -0.017  (-0.018, -0.016) | <0.001 | -0.022  (-0.024, -0.021) | <0.001 | -0.012  (-0.013, -0.011) | <0.001 | -0.009  (-0.010, -0.008) | <0.001 | -0.006  (-0.007, -0.005) | <0.001 |
| Intercept of control group | 0.173  (0.057, 0.290) | 0.004 | 0.371  (0.252, 0.490) | <0.001 | 0.246  (0.133, 0.360) | <0.001 | 0.123  (0.013, 0.232) | 0.0277 | 0.214  (0.096, 0.333) | <0.001 |
| Difference in baseline | 0.059  (-0.018, 0.136) | 0.132 | 0.034  (-0.057, 0.126) | 0.464 | 0.014  (-0.077, 0.105) | 0.762 | 0.033  (-0.043, 0.110) | 0.395 | 0.040  (-0.047, 0.127) | 0.363 |
| Slope of control group | -0.037  (-0.039, -0.034) | <0.001 | -0.008  (-0.011, -0.004) | <0.001 | -0.038  (-0.042, -0.035) | <0.001 | -0.036  (-0.039, -0.034) | <0.001 | -0.053  (-0.056, -0.050) | <0.001 |
| Difference in slope before MI | 0.003  (-0.023, 0.028) | 0.835 | -0.003  (-0.033, 0.028) | 0.869 | 0.013  (-0.018, 0.045) | 0.407 | -0.006  (-0.032, 0.021) | 0.679 | 0.022  (-0.006, 0.050) | 0.126 |
| Acute cognitive change after MI | 0.022  (-0.071, 0.116) | 0.640 | 0.038  (-0.075, 0.151) | 0.509 | -0.033  (-0.159, 0.092) | 0.605 | 0.005  (-0.099, 0.109) | 0·928 | 0.023  (-0.091, 0.136) | 0.696 |
| Changes in slope after MI | -0.013  (-0.045, 0.019) | 0.432 | -0.001  (-0.039, 0.037) | 0.957 | -0.024  (-0.065, 0.018) | 0.267 | 0.007  (-0.025, 0.040) | 0.x655 | -0.028  (-0.064, 0.007) | 0.117 |
| -2log likelihood | 82831.1 |  | 101177.2 |  | 104300.2 |  | 99846.2 |  | 107262.8 |  |

^a^Adjusted for baseline age, sex, educational level, marital status, area of residence, smoking, drinking, number of instrumental activities of daily living (IADLs), depression, hypertension, diabetes, high total cholesterol, lung diseases, and cancer.

^b^All cognitive values are z-score transformed.

#### 3.5 Supplementary Table 5. Trajectories of cognitive z scores among all participants over time after adjusting for BMI (421 in the MI group and 11287 in the control group)^a,b^

|  | Global cognition | | Episodic Memory |  | Visuospatial ability |  | Orientation |  | Attention and calculation |  |
| --- | --- | --- | --- | --- | --- | --- | --- | --- | --- | --- |
|  | β (95% CI) | *p* | β (95% CI) | *p* | β (95% CI) | *p* | β (95% CI) | *p* | β (95% CI) | *p* |
| Variables |  |  |  |  |  |  |  |  |  |  |
| Baseline age | -0.017  (-0.019, -0.015) | <0.001 | -0.023  (-0.025, -0.021) | <0.001 | -0.012  (-0.013, -0.010) | <0.001 | -0.010  (-0.011, -0.008) | <0.001 | -0.006  (-0.008, -0.005) | <0.001 |
| Intercept of control group | -0.116  (-0.285, 0.054) | 0.181 | 0.209  (0.039, 0.379) | 0.159 | 0.097  (-0.068, 0.261) | 0.249 | -0.109  (-0.266, 0.047) | 0.172 | 0.080  (-0.091, 0.250) | 0.361 |
| Difference in baseline | 0.069  (-0.020, 0.158) | 0.130 | 0.017  (-0.089, 0.123) | 0.750 | 0.006  (-0.100, 0.111) | 0.917 | 0.041  (-0.047, 0.128) | 0.360 | 0.051  (-0.049, 0.152) | 0.317 |
| Slope of control group | -0.038  (-0.040, -0.035) | <0.001 | -0.010  (-0.014, -0.006) | <0.001 | -0.039  (-0.043, -0.035) | <0.001 | -0.037  (-0.040, -0.035) | <0.001 | -0.053  (-0.056, -0.050) | <0.001 |
| Difference in slope before MI | -0.005  (-0.035, 0.024) | 0.722 | -0.004  (-0.039, 0.031) | 0.824 | 0.011  (-0.025, 0.047) | 0.541 | -0.004  (-0.034, 0.026) | 0.783 | 0.021  (-0.011, 0.054) | 0.201 |
| Acute cognitive change after MI | 0.027  (-0.081, 0.136) | 0.620 | 0.072  (-0.058, 0.202) | 0.279 | -0.067  (-0.212, 0.079) | 0.370 | 0.006  (-0.112, 0.123) | 0.925 | 0.028  (-0.101, 0.158) | 0.667 |
| Changes in slope after MI | -0.008  (-0.045, 0.030) | 0.681 | -0.013  (-0.059, 0.032) | 0.564 | -0.012  (-0.061, 0.037) | 0.621 | -0.002  (-0.040, 0.036) | 0.909 | -0.039  (-0.080, 0.002) | 0.065 |
| -2log likelihood | 63113.8 |  | 76822.4 |  | 79815.1 |  | 73953.8 |  | 80693.5 |  |

^a^Adjusted for baseline age, sex, educational level, marital status, area of residence, smoking, drinking, number of instrumental activities of daily living (IADLs), depression, BMI, hypertension, diabetes, high total cholesterol, lung diseases, and cancer.

^b^All cognitive values are z-score transformed.

#### 3.6 Supplementary Table 6. Trajectories of cognitive z scores among all participants over time only including participants who received cognitive tests in all four waves (270 in the MI group and 6318 in the control group)^a,b^

|  | Global cognition |  | Episodic Memory |  | Visuospatial ability |  | Orientation |  | Attention and calculation |  |
| --- | --- | --- | --- | --- | --- | --- | --- | --- | --- | --- |
|  | β (95% CI) | *p* | β (95% CI) | *p* | β (95% CI) | *p* | β (95% CI) | *p* | β (95% CI) | *p* |
| Variables |  |  |  |  |  |  |  |  |  |  |
| Baseline age | -0.012  (-0.014, -0.010) | <0.001 | -0.020  (-0.022, -0.018) | <0.001 | -0.009  (-0.011, -0.007) | <0.001 | -0.003  (-0.005, -0.001) | <0.001 | -0.002  (-0.004, 0.000) | 0.067 |
| Intercept of control group | 0.001  (-0.155, 0.156) | 0.991 | 0.308  (0.150, 0.467) | <0.001 | 0.183  (0.032, 0.335) | 0.018 | -0.152  (-0.289, -0.016) | 0.029 | 0.045  (-0.116, 0.207) | 0.583 |
| Difference in baseline | 0.088  (-0.007, 0.184) | 0.700 | 0.040  (-0.075, 0.155) | 0.493 | 0.021  (-0.094, 0.136) | 0.720 | 0.035  (-0.054, 0.124) | 0.444 | 0.117  (0.006, 0.228) | 0.038 |
| Slope of control group | -0.035  (-0.038, -0.032) | <0.001 | -0.004  (-0.008, 0.000) | 0.032 | -0.039  (-0.043, -0.035) | <0.001 | -0.020  (-0.022, -0.017) | <0.001 | -0.044  (-0.047, -0.041) | <0.001 |
| Difference in slope before MI | -0.005  (-0.035, 0.025) | 0.740 | -0.007  (-0.044, 0.030) | 0.715 | 0.013  (-0.025, 0.051) | 0.491 | -0.012  (-0.039, 0.016) | 0.407 | 0.003  (-0.032, 0.037) | 0.876 |
| Acute cognitive change after MI | 0.053  (-0.061, 0.168) | 0.361 | 0.064  (-0.075, 0.204) | 0.365 | -0.035  (-0.193, 0.123) | 0.667 | 0.032  (-0.084, 0.148) | 0.583 | 0.087  (-0.056, 0.231) | 0.234 |
| Changes in slope after MI | -0.012  (-0.049, 0.025) | 0.518 | 0.001  (-0.044, 0.047) | 0.956 | -0.025  (-0.075, 0.025) | 0.329 | 0.004  (-0.029, 0.037) | 0.818 | -0.031  (-0.074, 0.011) | 0.149 |
| -2log likelihood | 51482.7 |  | 63989.2 |  | 65297.8 |  | 52094.1 |  | 63334.8 |  |

^a^Adjusted for baseline age, sex, educational level, marital status, area of residence, smoking, drinking, number of instrumental activities of daily living (IADLs), depression, hypertension, diabetes, high total cholesterol, lung diseases, and cancer.

^b^All cognitive values are z-score transformed.
